# Deviations in Predicted COVID-19 cases in the US during early months of 2021 relate to rise in B.1.526 and its family of variants

**DOI:** 10.1101/2021.12.06.21267388

**Authors:** P. Nandakishore, M. Liu, Prakash R., S. Gourneni, R. Sukumaran, J.M. Berman, S. Iyer, C. Glorioso

## Abstract

**Objective:** To investigate the abrogation of COVID-19 case declines from predicted rates in the US in relationship to viral variants and mutations.

**Design:** Epidemiological prediction and time series study of COVID-19 in the US by State.

**Setting:** Community testing and sequencing of COVID-19 in the US.

**Participants:** Time series US COVID-19 case data from the Johns Hopkins University CSSE database. Time series US Variant and Mutation data from the GISAID database.

**Main outcome measures:** Primary outcomes were statistical modeling of US state deviations from epidemiological predictions, percentage of COVID-19 variants, percentage of COVID-19 mutations, and reported SARS-CoV-2 infections.

**Results:** Deviations in epidemiological predictions of COVID-19 case declines in the North Eastern US in March 2021 were highly positively related to percentage of B.1.526 (Iota) lineage (*p <* 10*e* − 7) and B.1.526.2 (*p <* 10 − 8) and the T95I mutation (*p <* 10*e* − 9). They were related inversely to B.1.427 and B.1.429 (Epsilon) and there was a trend for association with B.1.1.7 (Alpha) lineage.

**Conclusion:** Deviations from accurate predictive models are useful for investigating potential immune escape of COVID-19 variants at the population level. The B.1.526 and B.1.526.2 lineages likely have a high potential for immune escape and should be designated as variants of concern. The T95I mutation which is present in the B.1.526, B.1.526.2, and B.1.617.2 (Delta) lineages in the US warrants further investigation as a mutation of concern.

## Introduction

Since the middle of 2020 many variants of SARS-CoV-2 have emerged. The emergence of these variants has led to second and third waves of COVID-19 cases in many countries including the US, Britain, and India. These variants have claimed numerous lives and are responsible for the overcrowding of hospitals and other healthcare services. Each SARS-CoV-2 variant is characterized by mutations on the genome of SARS-CoV-2. Some of these mutations impart advantages to the variants such as higher transmission rates or escape from immunity from previous COVID-19 infection or vaccination.

Many epidemiological models predicted a decline in COVID-19 cases in the US in February and March of 2021 from a combination of natural immunity and previous infection [1]. Epidemiologists expected that this may be delayed by variants of concern [1]. The B.1.1.7 (Alpha) lineage (UK origin) was amongst the most concerning of the known variants in the US. The B.1.1.7 lineage was found to be 43 to 90% more transmissible compared to the previous variants based on the use of social contact, mobility data, and other demographic indicators of COVID-19 [2]. Consequently, studies point out that the number of cases would be considerably higher compared to what was observed in 2020, without fast-paced vaccine roll-out [1] [2](5,6). The B.1.1.7 variant (UK origin) had a significant decrease in neutralization, specifically among populations aged above 55 [3](9). The B.1.1.7 variant does not appear to raise concerns regarding the vaccine’s efficacy in real world or antibody neutralization studies [4] [5](7,8). The US also had two variants that originated in California-B.1.429 and B.1.427 (Epsilon). These exhibit a 18.6-24% increase in transmission compared to wild-type. B1.427 [6] specifically shows a 3-6 fold reduction in neutralizing titers of vaccinated or convalescent individuals [7]. It can be expected that so long as transmission rates are high, new variants will continue to emerge, and each must be treated as a new threat with the potential to escape prior immunity.

Other variants of concern, which included the P.1 and P.2 (Zeta) lineages (Brazilian origin) and the B.1.351 (Beta, S. African origin) were not in the US in significant enough numbers to pose an immediate threat. Although, the B.1.351 variant has shown potential for immune escape and therefore can reduce the efficiency of vaccines [3](9).

Despite the CDC and other epidemiological models predicting case declines in late 2020 and early 2021, the US saw continued cases in the winter and spring of 2021 in the US with New York City as the epicenter (fig 1A, 5). In addition, a new variant arising from the Washington Heights area of New York was identified([8]).

**Figure 1.**
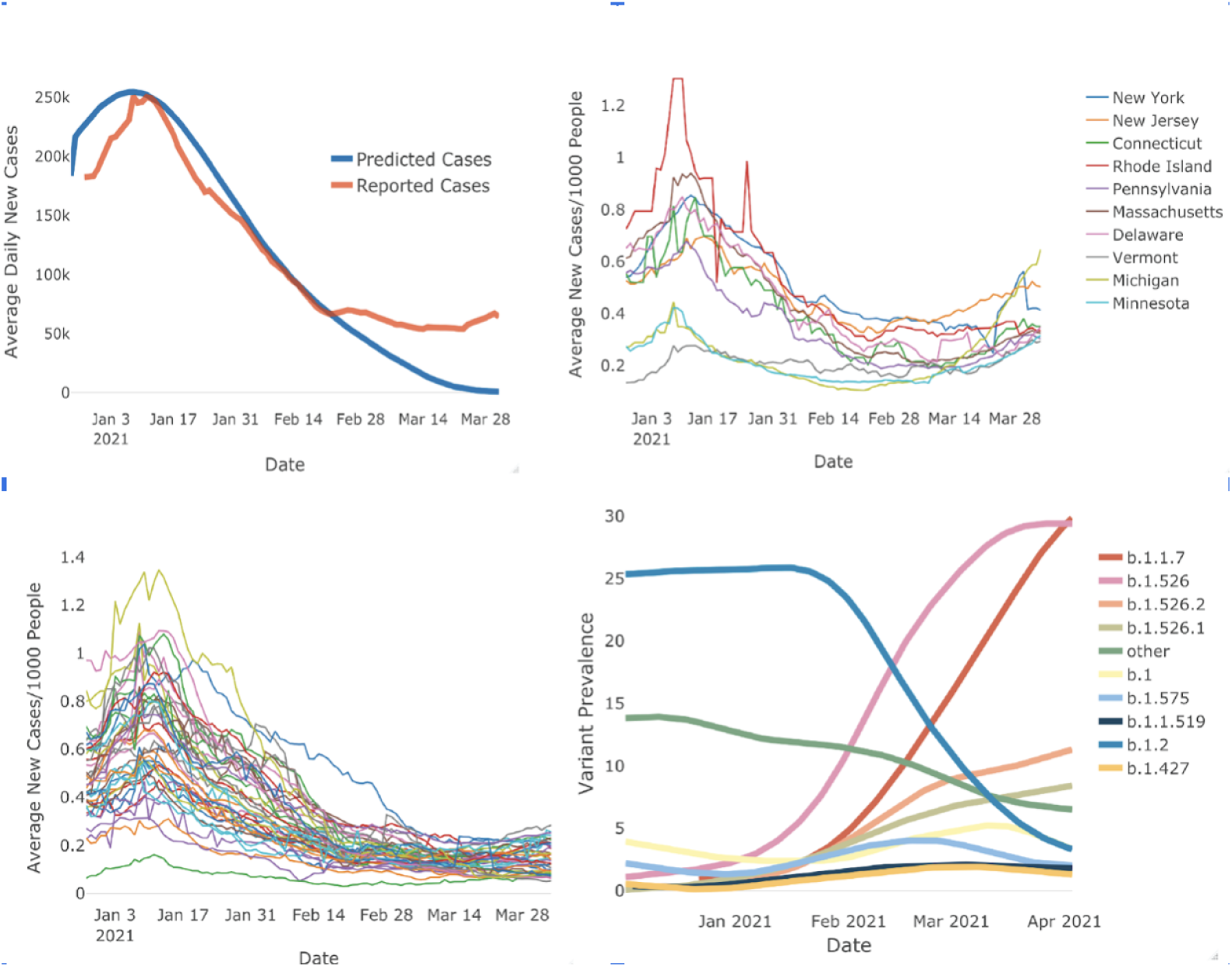
Rise of new B.1.1.7 and b.1.526 variants in Northeast US coincides with a rise in COVID-19 cases. Panel A (Top left) shows the SIR Model for predicting daily COVID-19 cases. The blue curve represents the model, the orange curve represents the observed case numbers. The model follows the observed case numbers closely however diverges around end of February. Panel B (Top left) shows daily case numbers per 1000 people for the states in the Northeastern US. daily case numbers fall around mid-February but start increasing by the month end. This trend is absent in the rest of the US States as shown in Panel c (Bottom Left). Panel D (Bottom right) Shows the time series of variant prevalence in New York State. Over January and February, B.2 which is the original COVID-19 strain decreases in prevalence but we observe that B.1.1.7 and B.1.526 increase in prevalence.

We demonstrate that the predicted drop in cases in the US in February and March of 2021 was delayed predominantly by infections in the North Eastern US by variants that originated in New York City, B.1.526 and B.1.526.2 [9]. We first explore the relationship between the number of COVID-19 cases and prevalence of various variants and mutations. We show that there is a positive correlation between the number of cases and the prevalence of the B.1.526 family of variants. The prevalence of mutations that define the variant B.1.526 also shows a positive correlation with the number of cases over time.

## Methods

### 0.1 Data resources

The COVID-19 variants data was sourced from the API of Outbreak.info [10]. Outbreak.info aggregates data across scientific sources, though the API, daily surveillance reports about lineages and mutations, countries, states, and counties from GISAID Initiative [11] were extracted for analysis. In an effort to include the maximum number of lineages, the data [12] period included January 1st, 2021, through March 31th, 2021. COVID-19 cases and deaths come from dashboard Coronavirus Visual Dashboard operated by the Johns Hopkins University Center for Systems Science and Engineering (JHU CSSE) [12]. The state population origins from census data of the United States Census Bureau in 2019 [13].

### 0.2 Data Pre-processing

#### 0.2.1 COVID-19 Variants and Mutation data

Each data point in figure 2 and figure 3 represents the average number of cases per thousand people plotted against the prevalence of a variant/mutation for a single state. Everything stated below applies to both the variant data in fig 2 and mutation data in fig 3. Prevalence is defined as the number of cases of a variant COVID-19 divided by the number of recorded COVID-19 cases on a day. The outbreak.info API provides a prevalence measure for each day hence the prevalence was averaged across all days in a month. Prevalence data for each month was collected for all states then averaged for each state over a month. Gaussian smoothing with a window of 7 days applied to the time series of the variant data. The Gaussian smoothing reduced the effect of noise due to spikes in the time series data. Varying the window for Gaussian smoothing from 3 to 10 days did not significantly alter the results. These spikes represent cumulative variant case number data dumps on specific days hence they were a source of noise while averaging prevalence over a single month.

**Figure 2.**
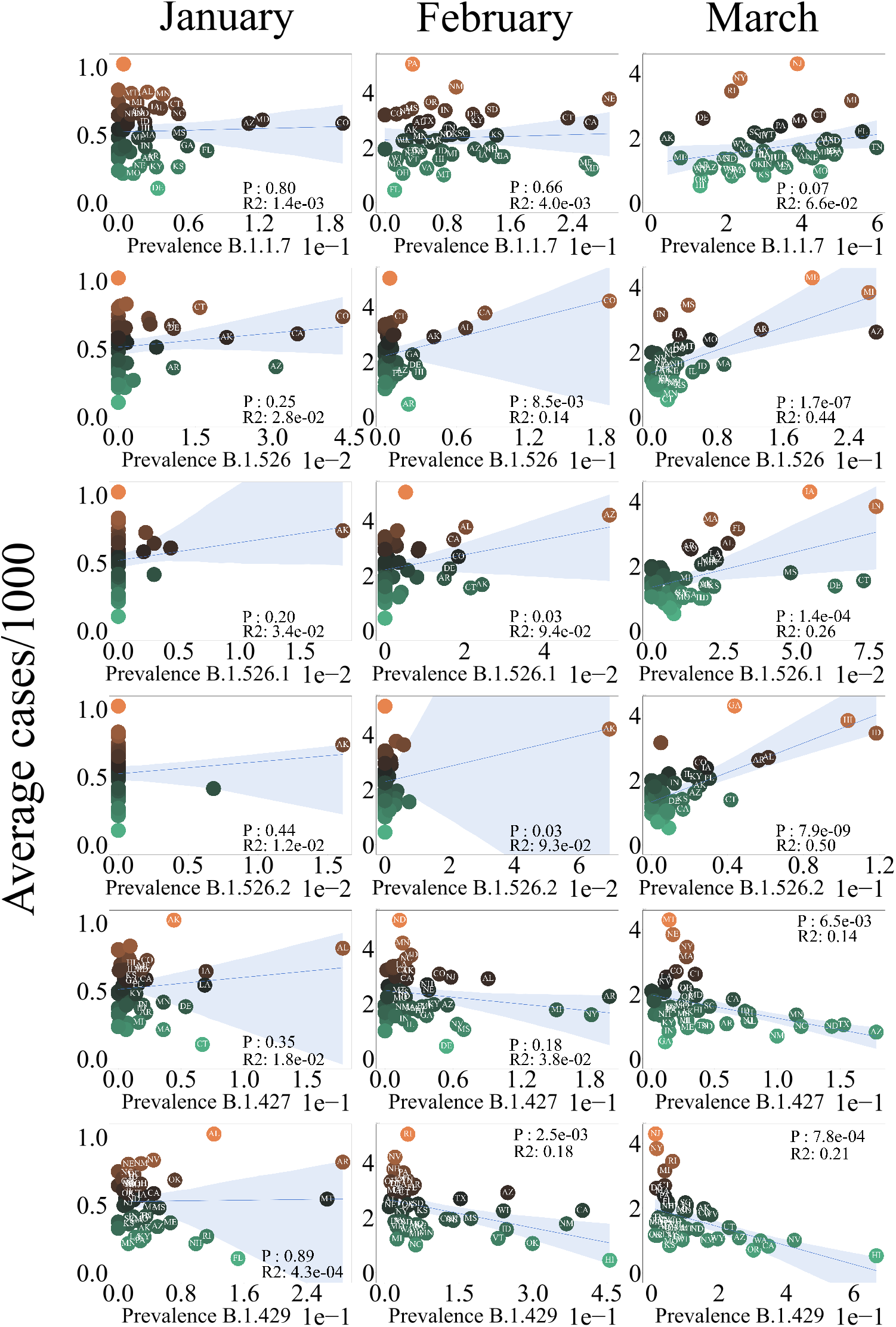
OLS Regression analysis of average number of cases per thousand vs COVID-19 lineage prevalence. Right column represents regression analysis for January, center for February and left for March. States that have a prevalence of larger than 0.01 are labeled. The shaded blue region represents the error in the slope of the fit. The color each each data points represents the scale of the average number of cases.

**Figure 3.**
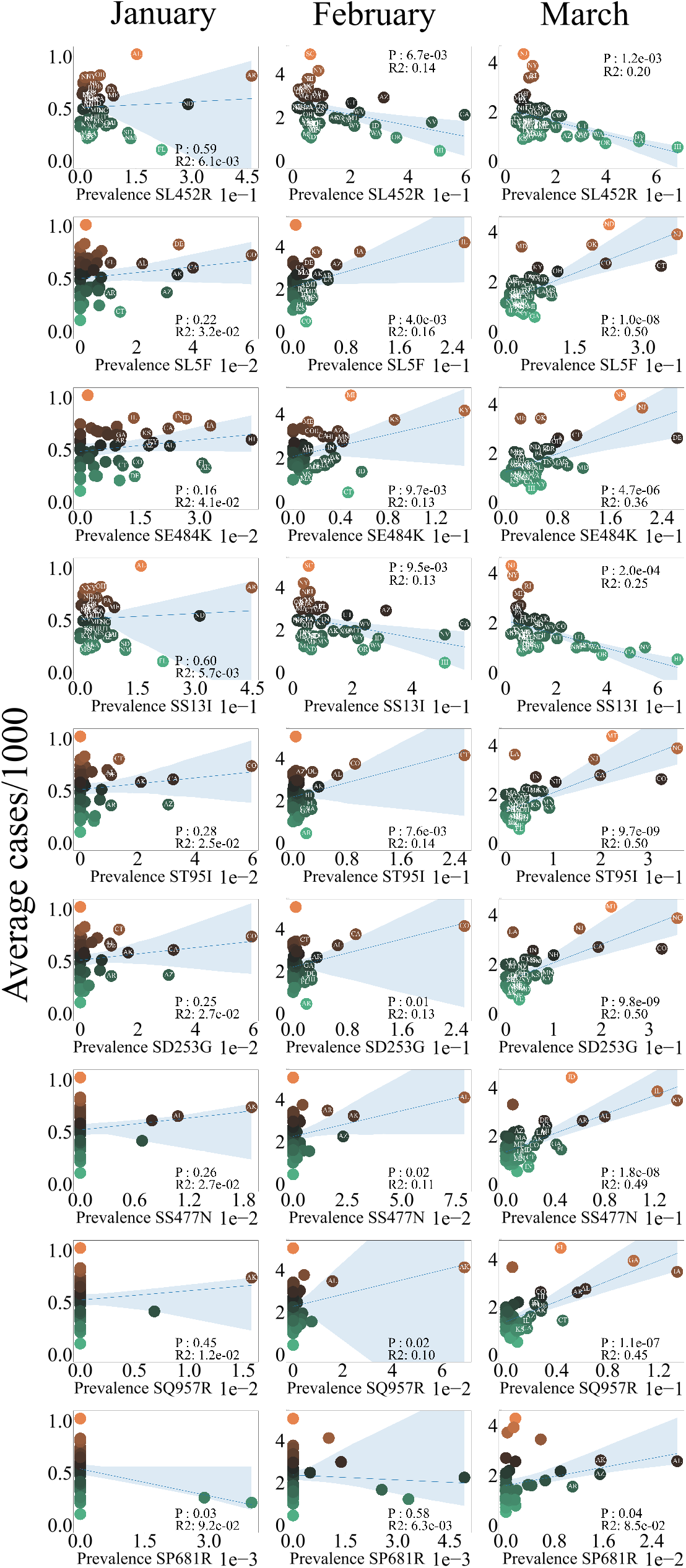
OLS Regression analysis of average number of cases per thousand vs COVID-19 Mutations. All mutations that are significant over January, February and March are plotted.

**Figure 4.**
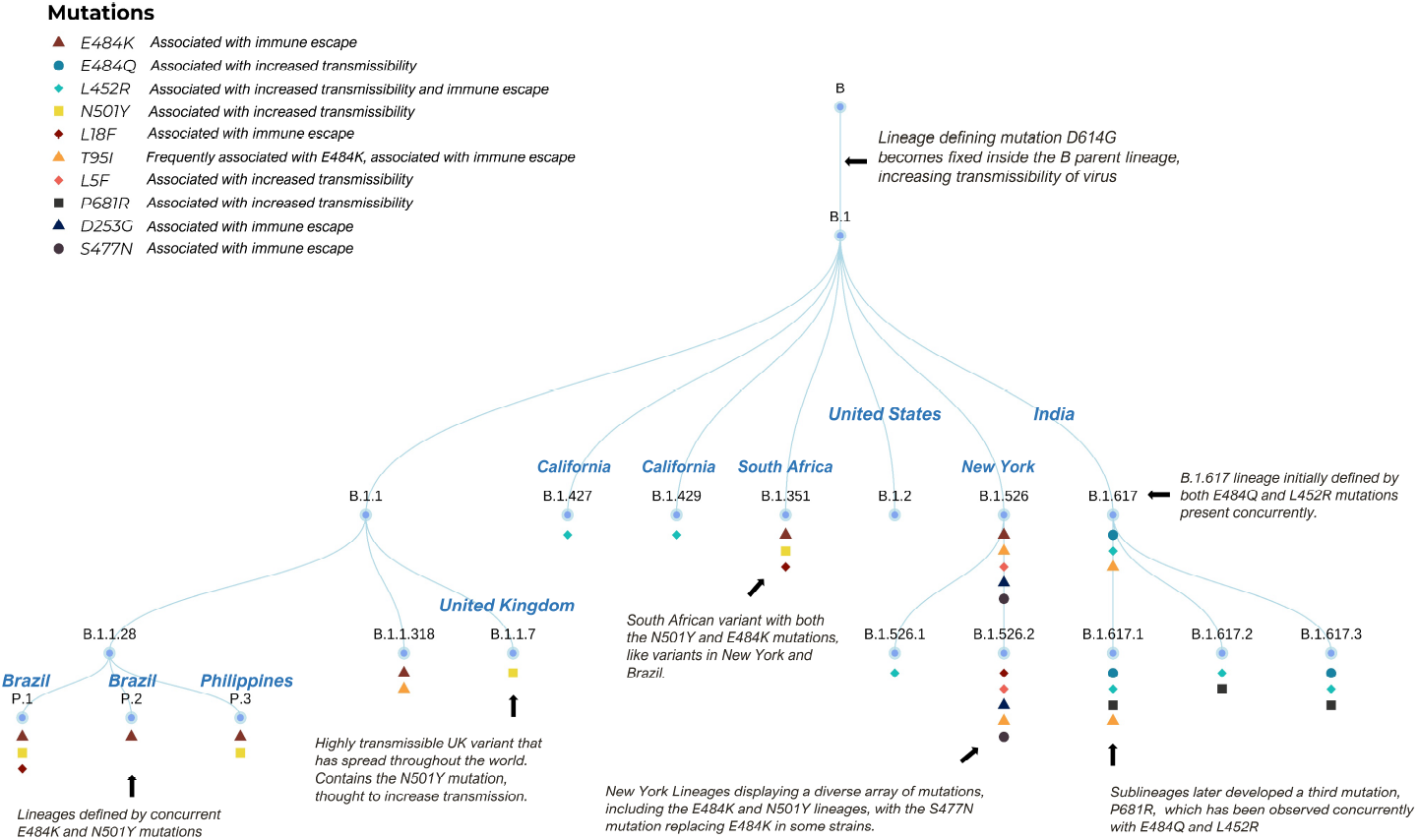
A Dendrogram outlining the relationship between COVID-19 lineages. The top right inset lists the definitions of each symbol.

The ordinate is scaled close to 1 by dividing the number of COVID-19 cases per day by the population of the state divided by a thousand. The order of magnitude of COVID-19 cases had decreased almost by an order of magnitude post the second wave of cases in December 2020 hence scaling the cases per day by state population per thousand negated the disparity between the most and least populous states. Python’s Pandas [14] package is used to process the data.

#### 0.2.2 Variants Map

For each state and each month, the top three frequent lineages were selected as the candidates for relative importance analysis based on the cumulative lineages cases. To standardize the COVID-19 cases and deaths, the COVID-19 case ratio was calculated by the COVID-19 cases of each state divided by the state population, and COVID-19 fatality ratio was presented by COVID-19 deaths of each state divided by the state COVID-19 cases.

### 0.3 Analysis Methods

#### 0.3.1 Hybrid Regression-SIR Model

We use linear regression to estimate initial susceptible/infectious/recovered (SIR) parameters. We solve SIR differential equations for the next day for each region, then re-estimate the SIR parameters with that day’s output and the lag-adjusted non-pharmaceutical interventions (NPIs). We then iterate each day until reaching the end date. Total US cases are calculated from the sum of the cases for each state. Holidays are adjusted for by addition to beta.

The algorithm has seven basic steps:

- Linear regression including NPIs from Oxford database to estimate the initial SIR parameter, *β* (infection-producing contacts per unit time).
- Adjust the parameters to account for asymptomatic cases based on CDC seroprevalence estimates [15].
- Employ a separate SIR model using the *β* estimated in step one for the next day for each US state.
- Feed the new *β* extracted from each SIR model into a linear regression model that incorporates the NPIs from one week prior (accounting for 14-day lag time from NPI to case reporting) to produce an NPI-adjusted *β*.
- Repeat steps 2-4 each day, feeding in the previous days predicted NPI-adjusted *β* until reaching the end date.
- For major holidays, input variables are adjusted up by adding the equivalent of three extra cases per 100,000 people per county, with delays to account for event-to-reporting lag time.
- The total US cases are summed from the US state cases.

The mathematical description of the ordinary differential equations (ODE) implemented are:

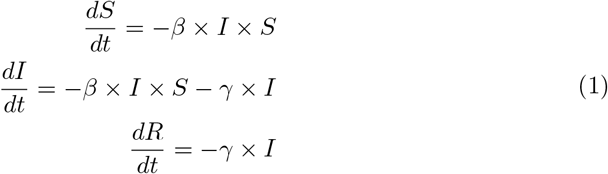

While SIR modeling is useful for accurate predictions, it is not sufficient, as it fails to consider that the reproductive number, *R*_0_ is not a constant and will vary by country, region, and over time, as conditions and interventions change. This dynamic *R*_0_ is often described as *R*_*e*_(*R*_*effective*_). Additionally, the number of susceptible, infectious, and recovered individuals also vary by region. Testing capacity and the relationship of reported cases to actual cases also varies by region. Finally, initial infection dates are regionally-specific, as SARS-CoV-2 originated in China, spread to other parts of Asia, then to Europe, to the Americas and so on. Together, these regional-specificities require that each region must be predicted separately with its own SIR model.

#### 0.3.2 Regression Analysis

Regression Analysis OLS regression of the number of cases per thousand was performed on the prevalence for numerous variants and mutations as shown in 2 and fig 3. The python package statsmodels [12](12) was used to perform the analysis. The p-values and R2 for each regression is provided in the inset of each figure.

#### 0.3.3 Relative Importance Analysis

Relative importance analysis is a useful technique to determine the relative importance of predictors when independent variables are correlated to each other. In our case, we hope to know how the most influential lineage changed regarding COVID-19 infections and deaths in each state. A relative importance analysis was performed using the “relaimpo” package in R [13]. Each predictor was fitted with a simple linear regression model to compare the R^2^-values, these uni-variate R^2^-values indicate what each predictor alone can explain the response. The importance ranking was determined by the uni-variate R^2^-values-values, the larger is the uni-variate R^2^-values, the more important is the predictor. Besides the R^2^-values, we also used the p-value of coefficient (*p <* 0.05) in each model to filter insignificant predictors before we rank the importance. Therefore, it is possible that no candidate is significant in the models. In this way, we can determine which lineage is the most influential regarding new COVID-19 cases or deaths for each state and each month. All statistical analyses were conducted using R version 4.0.1 (2020-06-06), and figures were produced using the ggplot2 package [16].

## 1 Results

The classical susceptible/infectious/recovered (SIR) model is a versatile tool to predict the spread of infectious diseases. SIR models coupled differential equations which are solved to forecast dynamics disease spread. As with any set of differential equations, SIR models require a set of initial conditions for numerically solving them.

We built an SIR model to predict the number of cases in the United States. We used US Center for Disease Control (CDC) antibody data to adjust for under-testing and under-reporting. We also adjusted for Holiday travel and event related spikes by systematically increasing beta. Figure 1A shows the observed number of cases and the predicted number of cases according to the SIR model.

Our model was highly accurate for predicting eight weeks out, with a MAE of 10%. This is half the error rate of any model or ensemble on the CDC dashboard and was able to predict for twice as far out (data not shown).

In late-February 2021, cases abruptly deviated from the predicted trajectory (Fig 1A). Most active US cases starting at that time were clustered in the North Eastern US on a gradient decreasing from New York City (Fig 1C). When we evaluated case deviations by State, we found that cases only deviated from predictions in these North Eastern US states (Fig 1B). Other states were almost uniformly following the trajectory predicted by our mathematical model (Fig 1C).

### 1.1 Variant Analysis

We investigated which variants were predominant in the states that deviated from our model and found that B.1.526 and B.1.526.2 became the predominant variants in New York and neighboring New Jersey in mid-February (Fig 1D), the same time that cases began to deviate from our predictions (Fig 1A).

To assess which lineages are influencing the changing case numbers in the US in January, February, and March of 2021, we performed linear regression and relative importance analysis. The variants B.1.526, B.1.526.1, and B.1.526.2 (NYC origins) had the most significant positive relationships to case numbers, p *<* 2e^−7^ and p *<* 8e^−9^, respectively for March Fig 2. These variants transition from being non-significant in January and February to being significant in March (Fig 2). B.1.526.1 was also significant (p *<* 2e^−4^)(2). We also observed that B.1.427 and B.1.429 lineages (California origin) had a significant(p *<* 3e − 3 and p *<* 8e − 4) but inverse relationship to the number of cases as seen in (2). These lineages are associated with smaller case numbers consistent with the rapidly declining cases in California and the Pacific Northwest (1C). Surprisingly, the prevalence of the B.1.1.7 variant (UK origin) did not show significance with rising case numbers (Fig 2). B.1.1.7 did, however, associate with new COVID-19 cases and deaths in the greatest number of states in March.

We examined most prevalent variants in various US states by relative importance analysis in figures 7,8,9, where 7 shows which variants dominated in January of 2021, 8 shows which variants dominated in February of 2021, and 9 shows which variants dominarted in March of 2021. While B.1.526 rose in the prevalence over time it did not become an influential variant of concern till March of 2021. B.1.1.7 and B.2 variants and the original COVID-19 lineage dominate the map of number of cases in February and March.

**Figure 5.**
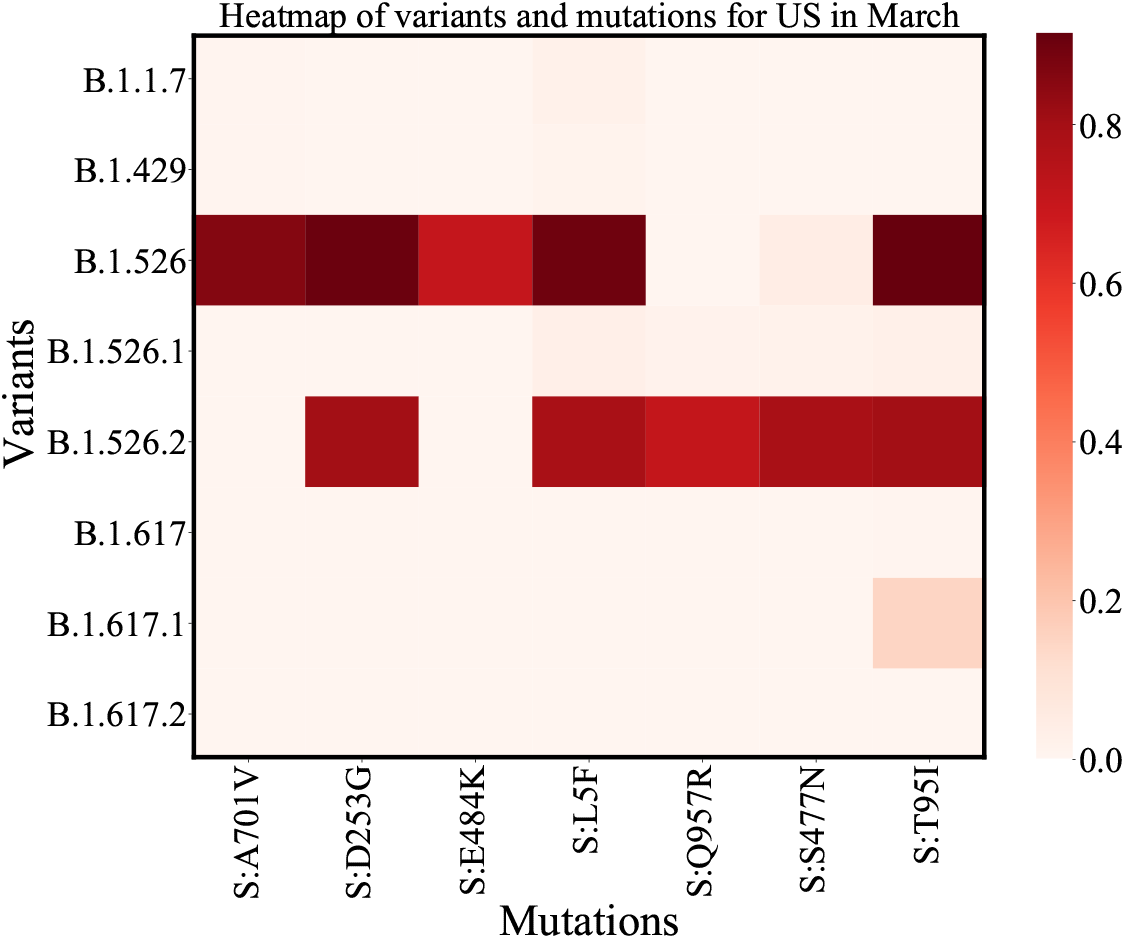
Heatmap for all US: Each cell in the heatmap represents the ratio of mutation-lineage case counts and lineage case counts. Data from January to the end of March has been used to generate the heatmaps. The Mutation-lineage count represents how many cases for a lineage contained a mutation. This ratio will always be less than 1 since the total number of mutation-lineage cases will be less than the total number of lineage cases. The darker shades in the heatmap represent higher counts of a mutation for a specific lineage.

**Figure 6.**
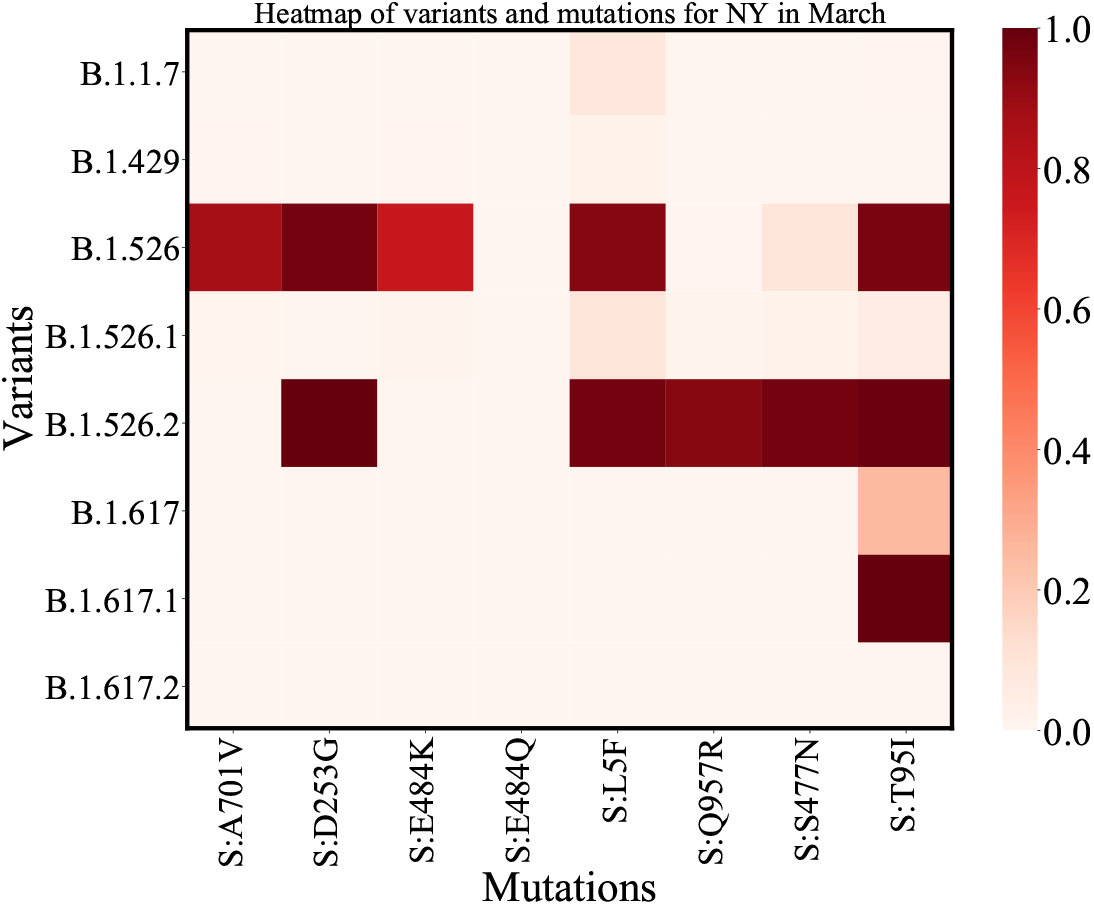
Heatmap for NY: Similar to the heatmap for all of US the darker regions represent higher mutation-lineage counts.

**Figure 7.**
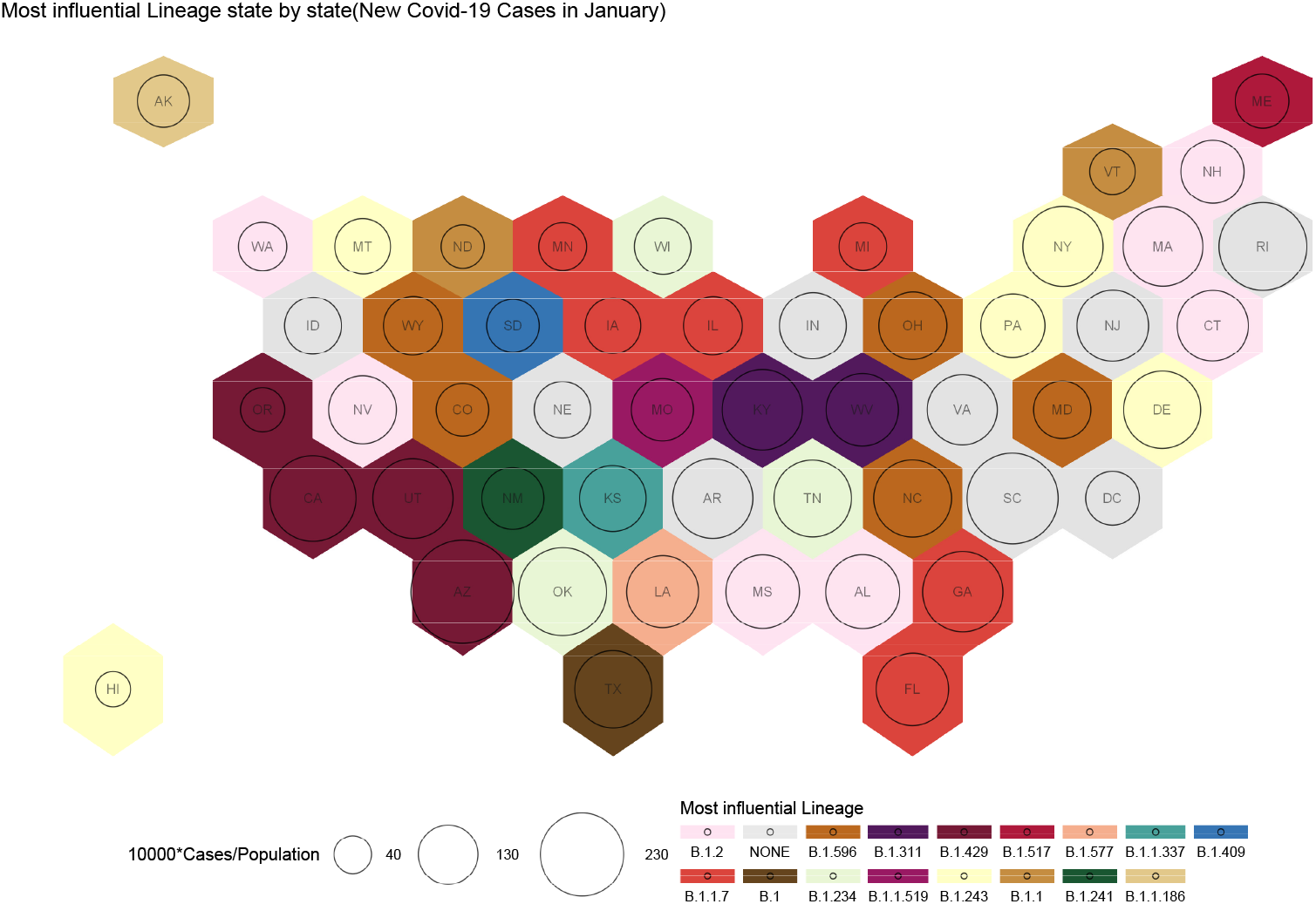
A map representing the most influence variants for the month of January. The circles represent cases in segments of 10000 as mentioned in the bottom left of the figure. The colors represent different variants. No single variant has emerged as an influential variant. The west coast has B.1.519 as the dominant variant and B.1.1.7 has its presence in six states. The None represents B.1 which is the original strain

**Figure 8.**
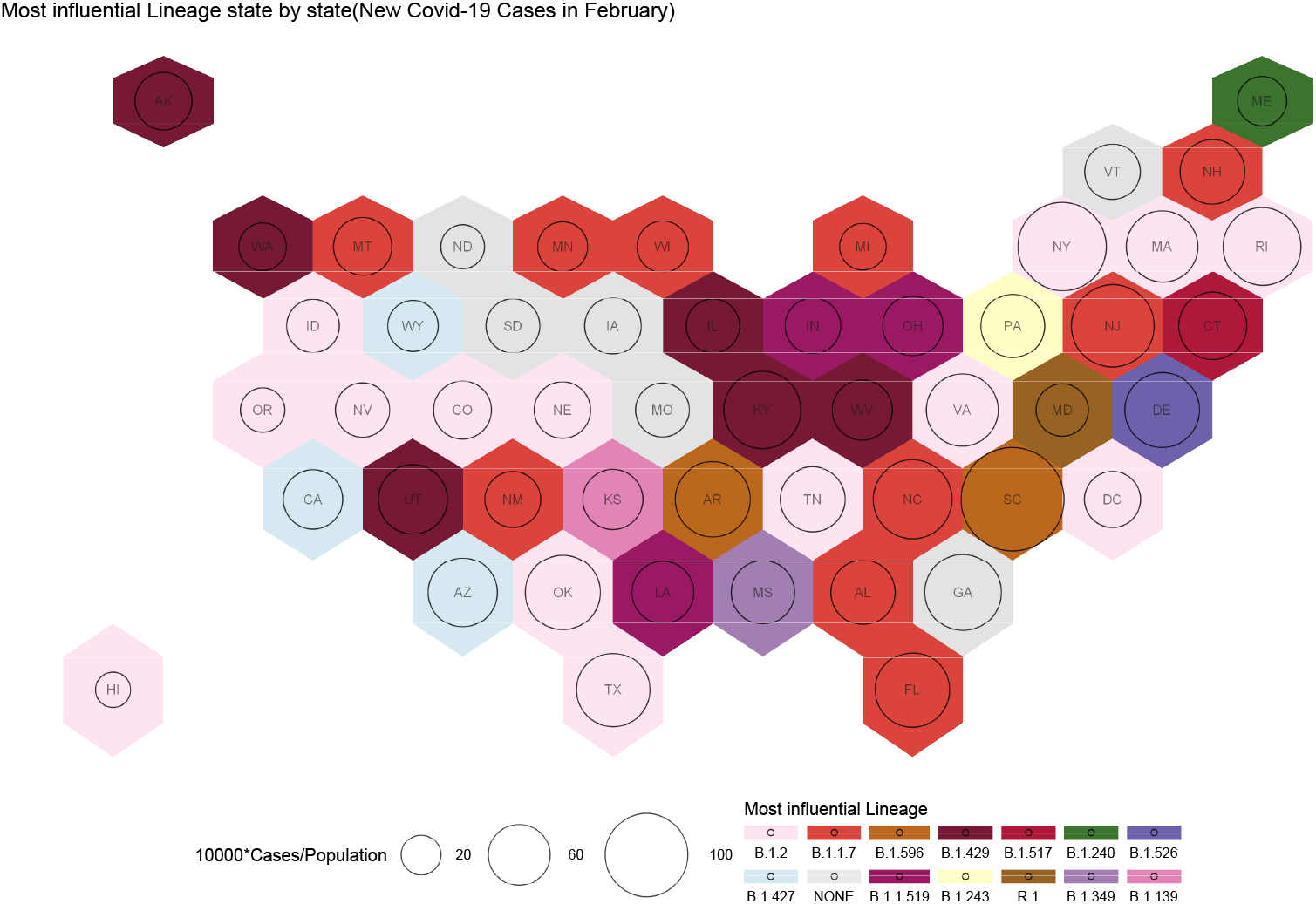
A map representing the most influential variants for the month of February. The circles represent cases in segments of 10000 as mentioned in the bottom left of the figure. The colors represent different variants. B.1.1.7 has spread to 10 States, B.1.427 has spreadf to three states in the western half of the US. B.1.526 emerges as the dominant variant in Delware

**Figure 9.**
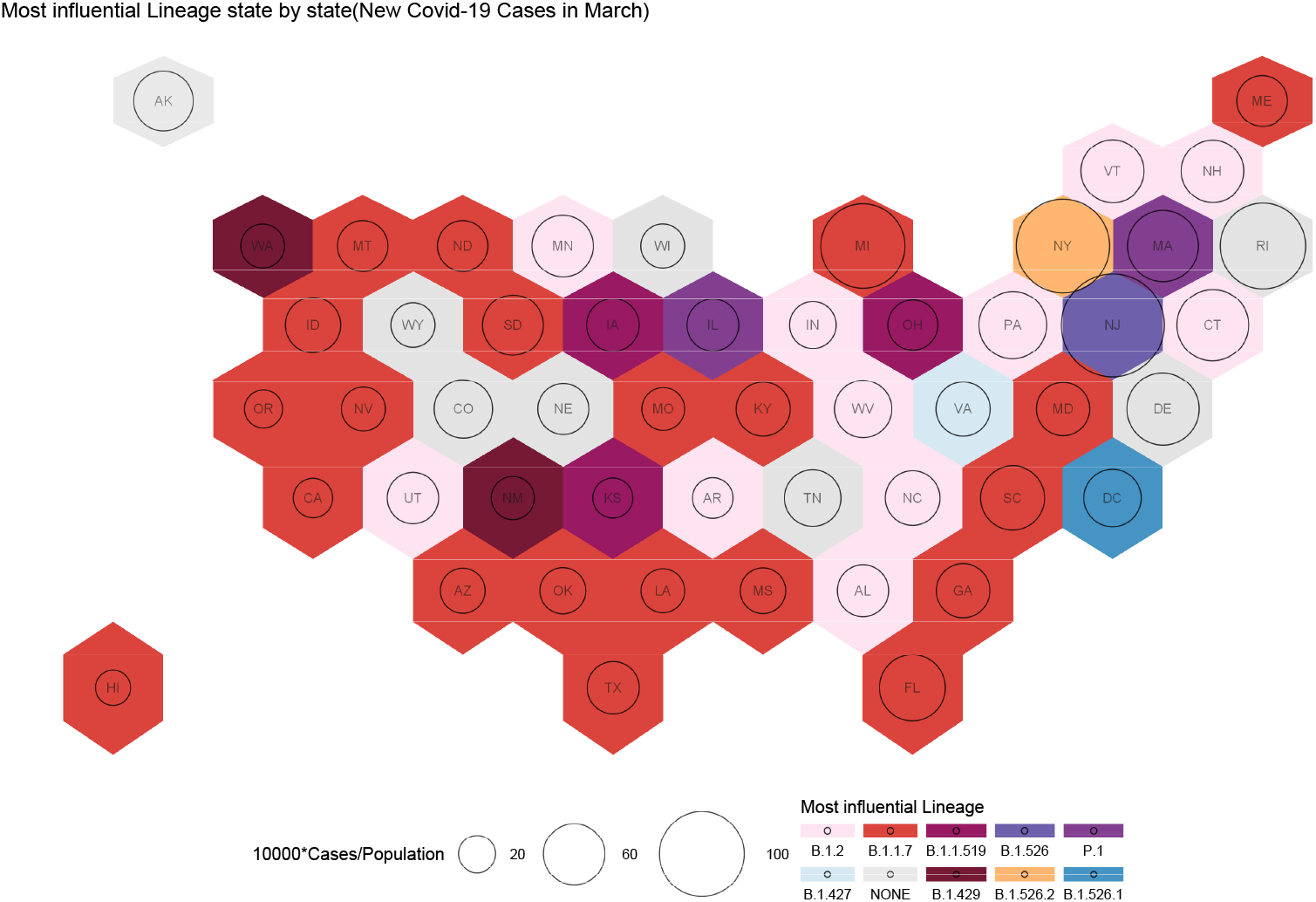
A map representing the most influential variants for the month of March. The circles represent cases in segments of 10000 as mentioned in the bottom left of the figure. The colors represent different variants. B.1.1.7 is the most influential variant in 21 states. B.1.2 is found in ten states. B.1.526 family of states are the most influential in three states in the North east - New York, New Jersey and DC

### 1.2 Mutation Analysis

These results led us to question how individual mutations affect the number of COVID-19 cases. We found that SL5F, SE848K, T951I, SD253G, SS477N, SQ957R and SP681R mutations showed a significant positively correlated relationship to the number of cases per 1000 (3). These mutations are a subset of the SARS-CoV-2 spike protein mutations that we explored, the p-value and *R*^2^ value for all the mutations we explore are presented in table 2. All mutations that were found to be positively correlated are associated with the B.1.526 family of variants (5), originating in New York. Interestingly, the much discussed SE848K “eek” mutation was not found in the B.1.526.2 or B.1.526.1 lineages but only in the parent variant B.1.526 as shown in figure 5. The mutations, SL452R and SS13I are also significant but were negatively correlated. These mutations are part of the B.1.427 and B.1.429 variants.

**Table 1.** P-value and R^2^ values for COVID-19-variants.

| Variant name | Month | P-val | $R^2$ |
| --- | --- | --- | --- |
| B.1.427 | January | 0.354 | 0.0179 |
| B.1.427 | February | 0.173 | 0.0383 |
| B.1.427 | March | 0.00653 | 0.1442 |
| B.1.429 | January | 0.889 | 0.0004 |
| B.1.429 | February | 0.00241 | 0.1762 |
| B.1.429 | March | 0.000779 | 0.2115 |
| B.1.1.7 | January | 0.797 | 0.0014 |
| B.1.1.7 | February | 0.662 | 0.004 |
| B.1.1.7 | March | 0.072 | 0.0658 |
| B.1.526 | January | 0.247 | 0.0278 |
| B.1.526 | February | 0.00841 | 0.136 |
| B.1.526 | March | 1.74e-07 | 0.437 |
| B.1.526.1 | January | 0.2 | 0.034 |
| B.1.526.1 | February | 0.0305 | 0.0939 |
| B.1.526.1 | March | 0.000138 | 0.2635 |
| B.1.526.2 | January | 0.441 | 0.0124 |
| B.1.526.2 | February | 0.0311 | 0.0932 |
| B.1.526.2 | March | 7.92e-09 | 0.5036 |
| B.1.351 | January | 0.175 | 0.038 |
| B.1.351 | February | 0.786 | 0.0016 |
| B.1.351 | March | 0.325 | 0.0202 |
| P.1 | January | 0.343 | 0.0187 |
| P.1 | February | 0.462 | 0.0113 |
| P.1 | March | 0.672 | 0.0038 |
| P.2 | January | 0.847 | 0.0008 |
| P.2 | February | 0.828 | 0.001 |
| P.2 | March | 0.761 | 0.002 |

**Table 2.**
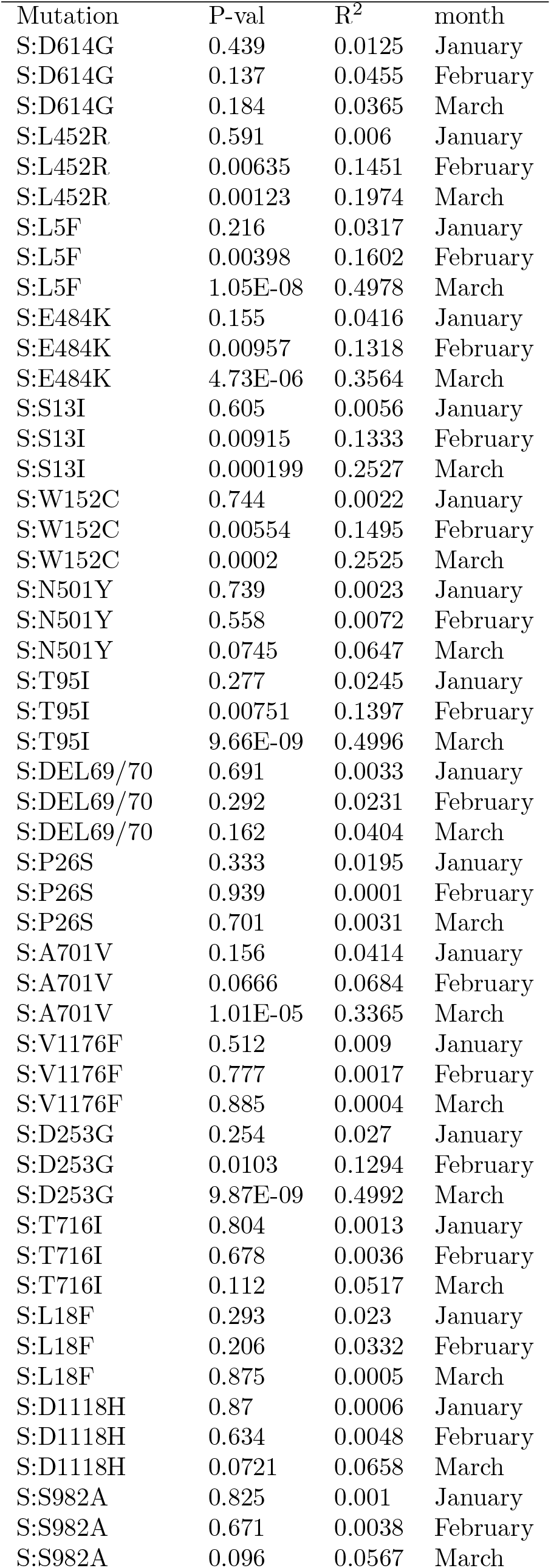
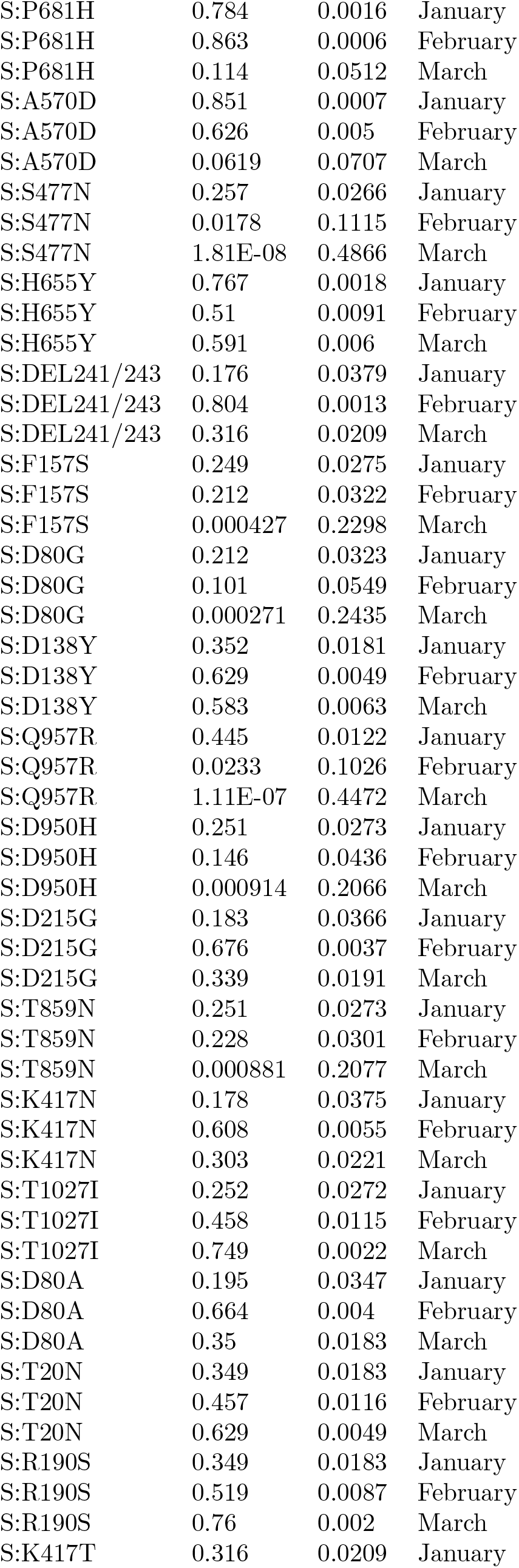
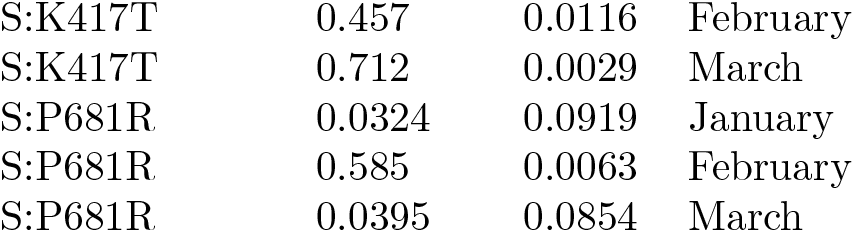
P-value and R^2^ values for COVID-19-mutations. We studied the correlation between the prevalence of multiple Spike mutations and number of cases and results presented in figure 3 is for a small subset of mutations.

| Mutation | P-val | R<sup>2</sup> | month |
| --- | --- | --- | --- |
| S:D614G | 0.439 | 0.0125 | January |
| S:D614G | 0.137 | 0.0455 | February |
| S:D614G | 0.184 | 0.0365 | March |
| S:L452R | 0.591 | 0.006 | January |
| S:L452R | 0.00635 | 0.1451 | February |
| S:L452R | 0.00123 | 0.1974 | March |
| S:L5F | 0.216 | 0.0317 | January |
| S:L5F | 0.00398 | 0.1602 | February |
| S:L5F | 1.05E-08 | 0.4978 | March |
| S:E484K | 0.155 | 0.0416 | January |
| S:E484K | 0.00957 | 0.1318 | February |
| S:E484K | 4.73E-06 | 0.3564 | March |
| S:S13I | 0.605 | 0.0056 | January |
| S:S13I | 0.00915 | 0.1333 | February |
| S:S13I | 0.000199 | 0.2527 | March |
| S:W152C | 0.744 | 0.0022 | January |
| S:W152C | 0.00554 | 0.1495 | February |
| S:W152C | 0.0002 | 0.2525 | March |
| S:N501Y | 0.739 | 0.0023 | January |
| S:N501Y | 0.558 | 0.0072 | February |
| S:N501Y | 0.0745 | 0.0647 | March |
| S:T95I | 0.277 | 0.0245 | January |
| S:T95I | 0.00751 | 0.1397 | February |
| S:T95I | 9.66E-09 | 0.4996 | March |
| S:DEL69/70 | 0.691 | 0.0033 | January |
| S:DEL69/70 | 0.292 | 0.0231 | February |
| S:DEL69/70 | 0.162 | 0.0404 | March |
| S:P26S | 0.333 | 0.0195 | January |
| S:P26S | 0.939 | 0.0001 | February |
| S:P26S | 0.701 | 0.0031 | March |
| S:A701V | 0.156 | 0.0414 | January |
| S:A701V | 0.0666 | 0.0684 | February |
| S:A701V | 1.01E-05 | 0.3365 | March |
| S:V1176F | 0.512 | 0.009 | January |
| S:V1176F | 0.777 | 0.0017 | February |
| S:V1176F | 0.885 | 0.0004 | March |
| S:D253G | 0.254 | 0.027 | January |
| S:D253G | 0.0103 | 0.1294 | February |
| S:D253G | 9.87E-09 | 0.4992 | March |
| S:T716I | 0.804 | 0.0013 | January |
| S:T716I | 0.678 | 0.0036 | February |
| S:T716I | 0.112 | 0.0517 | March |
| S:L18F | 0.293 | 0.023 | January |
| S:L18F | 0.206 | 0.0332 | February |
| S:L18F | 0.875 | 0.0005 | March |
| S:D1118H | 0.87 | 0.0006 | January |
| S:D1118H | 0.634 | 0.0048 | February |
| S:D1118H | 0.0721 | 0.0658 | March |
| S:S982A | 0.825 | 0.001 | January |
| S:S982A | 0.671 | 0.0038 | February |
| S:S982A | 0.096 | 0.0567 | March |
| S:P681H | 0.784 | 0.0016 | January |
| S:P681H | 0.863 | 0.0006 | February |
| S:P681H | 0.114 | 0.0512 | March |
| S:A570D | 0.851 | 0.0007 | January |
| S:A570D | 0.626 | 0.005 | February |
| S:A570D | 0.0619 | 0.0707 | March |
| S:S477N | 0.257 | 0.0266 | January |
| S:S477N | 0.0178 | 0.1115 | February |
| S:S477N | 1.81E-08 | 0.4866 | March |
| S:H655Y | 0.767 | 0.0018 | January |
| S:H655Y | 0.51 | 0.0091 | February |
| S:H655Y | 0.591 | 0.006 | March |
| S:DEL241/243 | 0.176 | 0.0379 | January |
| S:DEL241/243 | 0.804 | 0.0013 | February |
| S:DEL241/243 | 0.316 | 0.0209 | March |
| S:F157S | 0.249 | 0.0275 | January |
| S:F157S | 0.212 | 0.0322 | February |
| S:F157S | 0.000427 | 0.2298 | March |
| S:D80G | 0.212 | 0.0323 | January |
| S:D80G | 0.101 | 0.0549 | February |
| S:D80G | 0.000271 | 0.2435 | March |
| S:D138Y | 0.352 | 0.0181 | January |
| S:D138Y | 0.629 | 0.0049 | February |
| S:D138Y | 0.583 | 0.0063 | March |
| S:Q957R | 0.445 | 0.0122 | January |
| S:Q957R | 0.0233 | 0.1026 | February |
| S:Q957R | 1.11E-07 | 0.4472 | March |
| S:D950H | 0.251 | 0.0273 | January |
| S:D950H | 0.146 | 0.0436 | February |
| S:D950H | 0.000914 | 0.2066 | March |
| S:D215G | 0.183 | 0.0366 | January |
| S:D215G | 0.676 | 0.0037 | February |
| S:D215G | 0.339 | 0.0191 | March |
| S:T859N | 0.251 | 0.0273 | January |
| S:T859N | 0.228 | 0.0301 | February |
| S:T859N | 0.000881 | 0.2077 | March |
| S:K417N | 0.178 | 0.0375 | January |
| S:K417N | 0.608 | 0.0055 | February |
| S:K417N | 0.303 | 0.0221 | March |
| S:T1027I | 0.252 | 0.0272 | January |
| S:T1027I | 0.458 | 0.0115 | February |
| S:T1027I | 0.749 | 0.0022 | March |
| S:D80A | 0.195 | 0.0347 | January |
| S:D80A | 0.664 | 0.004 | February |
| S:D80A | 0.35 | 0.0183 | March |
| S:T20N | 0.349 | 0.0183 | January |
| S:T20N | 0.457 | 0.0116 | February |
| S:T20N | 0.629 | 0.0049 | March |
| S:R190S | 0.349 | 0.0183 | January |
| S:R190S | 0.519 | 0.0087 | February |
| S:R190S | 0.76 | 0.002 | March |
| S:K417T | 0.316 | 0.0209 | January |
| S:K417T | 0.457 | 0.0116 | February |
| S:K417T | 0.712 | 0.0029 | March |
| S:P681R | 0.0324 | 0.0919 | January |
| S:P681R | 0.585 | 0.0063 | February |
| S:P681R | 0.0395 | 0.0854 | March |

We also found that less than 30 cases of B.1.617 were present in the US during this time period and that both B.1.526 and B.1.526.2 variants share the T95I mutation with B.1.617. This observation is even more pronounced if we look at the heat map of mutations and variants for New York (6) where we observe the presence of ST95I in all family of B.1.526 variants.

### 1.3 The Variant Family tree

All strains of SARS-CoV-2 that we refer to as “variants” emerged from strain B.1. To better visualize this, we provide a dendrogram as shown in (4). The dendrogram documents variants in three geographical regions the US, UK and India. These regions have seen either an uptick in cases, as with India and the UK or a stalling in decline of COVID-19 cases (as in the the US). Specific mutations shared by certain strains of SARS-CoV-2 influence their virulence.

## 2 Discussion

The deviation in the number of cases in our epidemiological model shows the influence of variants on COVID-19 case numbers. Our results (figure 1C) show that B.1.526 had been rising in prevalence and led to the stall in fall of COVID-19 cases.

The increasing prevalence of B.1.526 may have been at least partially responsible for cases increases in many Northeastern states during February and March of 2021. This is demonstrated by the relationship between case counts and prevalence for B.1.526, B.1.526.1, and B.1.526.2, as discussed in results above.

Further support for this conclusion emerges from the growth of B.1.526 in New York and New Jersey as demonstrated in our relative importance analysis. By March, B.1.526 was the most variant causing the most deaths in New York, New Jersey and Connecticut, although B.1.1.7 was the dominant strain in the US overall.

Conversely, variants B.1.427 and B.1.429 had high prevalence prevalence in the Southern and Western part of the United States. The spread of these variants in these region is attributed to their geographical origin. Both these variants, as far as is known, originated in California. States like Nevada, Oregon and Arizona have significant number of cases of these variants.

Its also interesting to note that the variant B.1.427 does emerge as a dominant strain in the California, Wyoming and Arizona in February but is replaced by B.1.1.7 and the original COVID-19 strain in March. This observation fit with observation that B.1.427 has a negative correlation with the number of cases.

The relationship between mutation prevalence and number of cases follows a similar trend to the variant prevalence. Most of the mutations that become significant over time are associated with the B.1.526 family of variants.

These results have a broader implication beyond these variants. As new SARS-CoV-2 variants emerge, it remains possible that these new variants will escape immunity generated by infection with a prior variant, or acquired through vaccinations, defying the predictions of pandemic models, and leading to increased case-counts in regions with high expected immunity. Using regional deviations from highly accurate predictions can rapidly identify regions with variants that are escaping immunity.

### 2.1 Comparison with other works

The CDC observed both the emergence of B.1.526 in November 2020 [17] and that there was a sharp increase in the number of cases in New York by April 5^*th*^. This timeline is similar to other we have observed where by Mid-February B.1.526 prevalence had reached 30% prevalence by April 2020. The timeline is corroborated by Lasek-Nesselquist et al [8] as well who show detailed maps of the spread of B.1.526. They also mention that this variant has the E484K, S477N and D253G all of which are correlated with increase in number of COVID-19 in the US.

The CDC does mention that while B.152.6 does not cause more severe COVID-19 symptoms it could be more transmissible than other variants. This could explain its spread into other states and also its immune escape. As far as we know, we have not seen any other study document the relationship between the prevalence of COVID-19 variants and number of cases.

### 2.2 Limitations of the Study

The data used in this study are cross-sectional. Ideally we would be able to test whether the B.1.526 lineages were reinfecting the same person after being infected with the original strains. Additionally it would be ideal to have data on vaccination status and type of vaccine in the people infected with viral lineages longitudinally. Unfortunately, these data do not exist or are not accessible. We strongly encourage testing and reporting of this data by hospital systems so that we can more clearly understand the role of the variants on breakthrough cases. Another limitation of the study is delays in reporting sequencing data to databases. Accurate lineage data is available only with a three week delay. This means that we are delayed in understanding important information for policy planning and hospital staffing.

Our relative importance analysis while useful in understanding the most dominant lineages will fail to capture variants with rising prevalence or variants that dominate regionally. When a specific variant causes a large number of cases over a country it tends to be the focus of study. However the most dominant variant may not the most dangerous one, geographically located variants like B.1.526 are a cause for concern since they may escape notice due their limited presence.

### 2.3 Future Directions

In the future we hope to access longitudinal data to more directly assess the effect of lineages and their mutations on breakthrough cases and reinfection. These mutations need further study on their impact, particularly T95I. Lastly, we will soon hopefully be vaccinated to high enough degrees in more countries to take a global approach to looking at mutations, lineages, and vaccination. We also hope to map the biological significance of mutations to epidemiological outcomes. Currently there is limited understanding on what are the macro-effects of mutations hence this presents a challenge in accurately modelling the evolving pandemic.

### 2.4 Conclusion

Our article shows that the stalling of decline in COVID-19 cases in the US can be at least partially attributed to the rise of a geographically limited yet prevalent variant B.1.526. We come to this conclusion by studying the rise in prevalence of multiple variants and find that the B.1526 family of variants shows a positive correlation between the prevalence of the variants and the number the of cases per state. We initially observed a deviation using SIR modeling of the average number of cases in the first half 2021. This modeling showed that the number of predicted and actual cases differed from end of February of 2021. To better understand this deviation, we explored relationship between different variant prevalence and the average number of cases per state. We observed the positive correlations between the B.1.526 variant and the number of cases. This correlation between developed over the course of three months, January, February and March of 2021. We support this conclusion by also preforming relative importance analysis and showing that in many of the North-Eastern states B.1.526 family of variants are the most influential variants.

We have also explored the relationship between mutations and variant prevalence and found that the mutations present in the variants B.1.1.526 family of variants and B.1.1.7 seems to show a positive correlation with the number of cases in each state.

Newer variants will emerge as time progresses. Our hope is that the observations and analysis may help in understand how newer variants emerge from specific regions of the US and lead to more cases. Understanding which variants and mutational profiles are most concerning rapidly can aid in effectively designing policy for the global COVID-19 pandemic.

## Data Availability

All data produced in the present study are available upon reasonable request to the authors.

## Acknowledgments

We thank just about everybody.

### 3 Appendix

**Figure 10.**
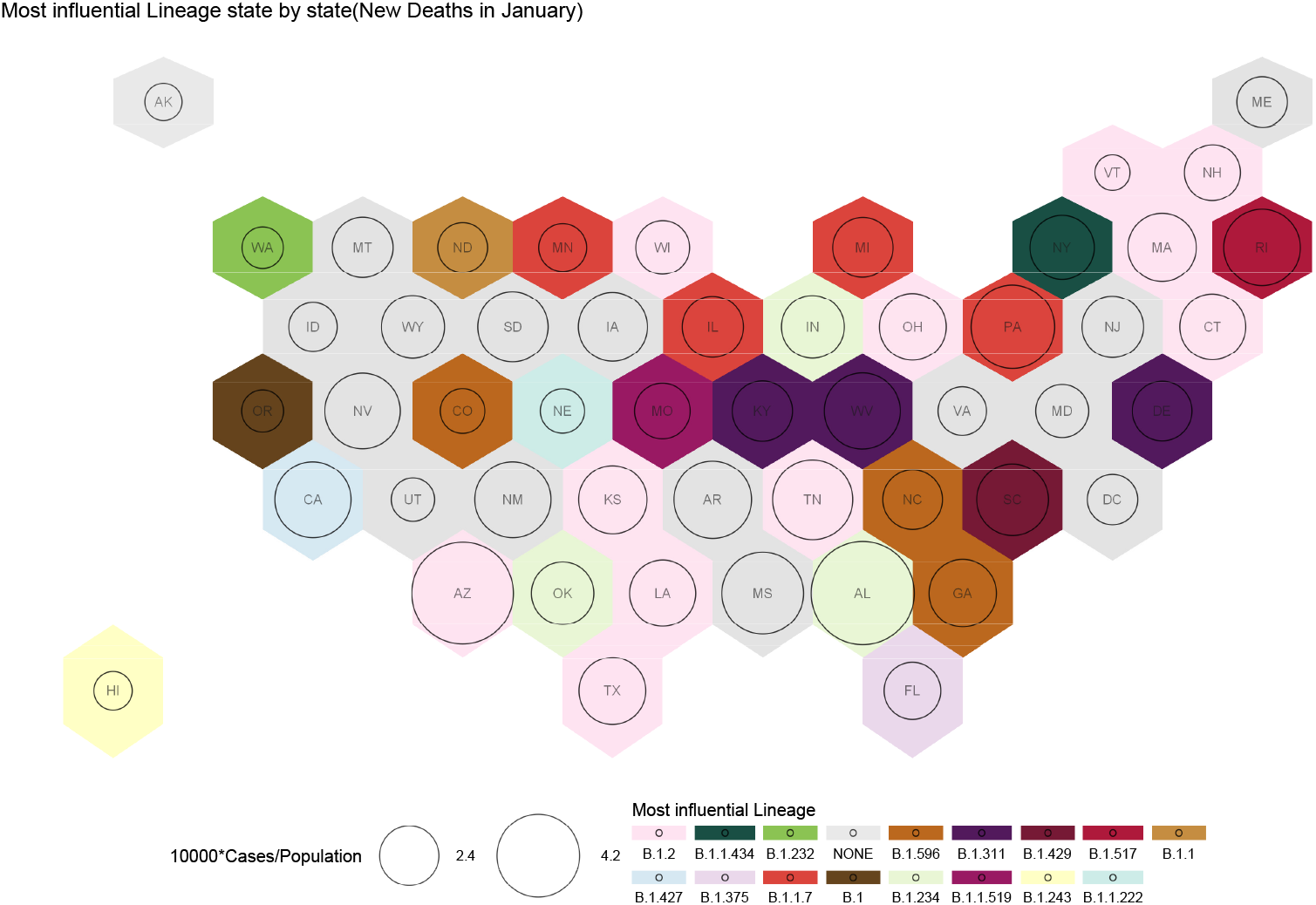
Influential variants causing deaths for the month of January. Each US State is presented as a hexagon. The circles represent cases in segments of 10000. The colors represent different variants. The none strain is Most of the deaths can be attributed to the original B.1 strain and the B.2 variant.

**Figure 11.**
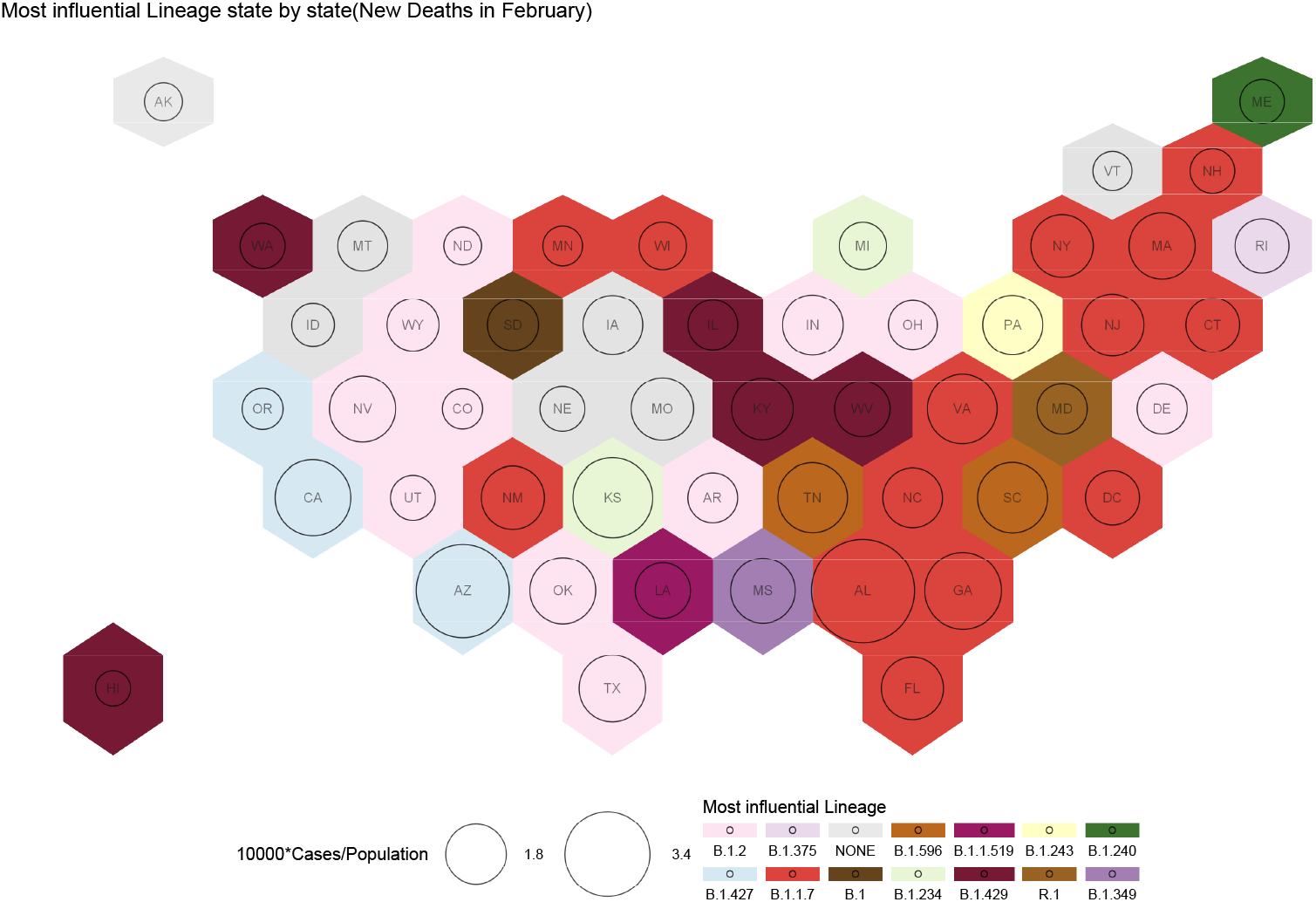
Influential variants causing deaths for the month of February. Each US State is presented as a hexagon. The circles represent cases in segments of 10000. The colors represent different variants. The deaths in the east coast is dominated by B.1.1.7 and in the west coast by B.1.427. B.2 is also responsible for deaths in the many of the mid-western states.

**Figure 12.**
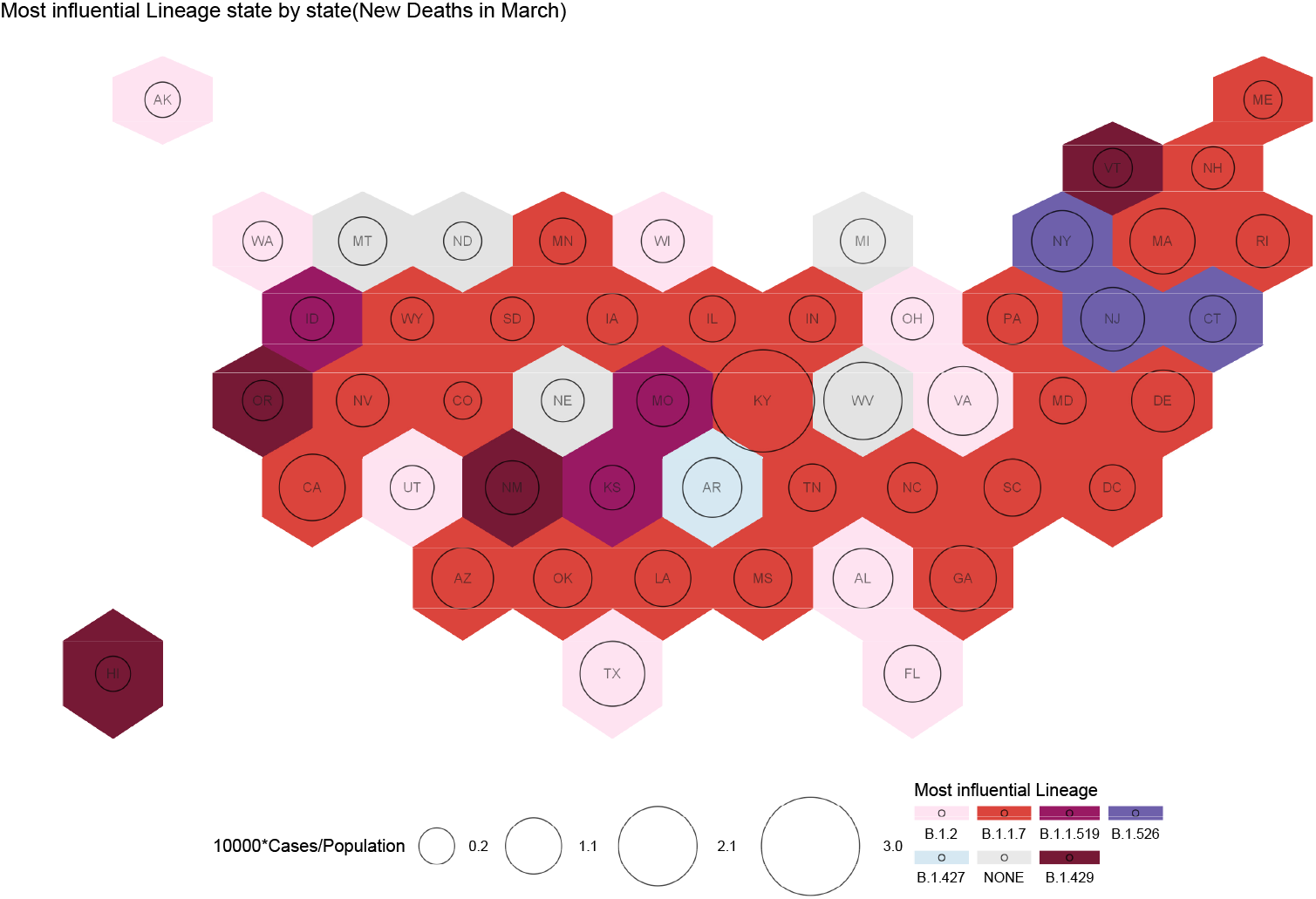
Influential variants causing deaths for the month of February. Each US State is presented as a hexagon. The circles represent cases in segments of 10000. The colors represent different variants. Most of the deaths by March are due to B.1.1.7. The B.1.526 family of variants cause the most number of deaths in the three states - New York, New Jersey and Connecticut. B.2 is responsible for deaths in 8 states spread out across the country

## References

1. Summer E. Galloway, P. Paul, D. MacCannell, Michael A. Johansson, J. Brooks, A. MacNeil, R. Slayton, S. Tong, B. Silk, G. Armstrong, M. Biggerstaff, and V. Dugan. Emergence of sars-cov-2 b.1.1.7 lineage united states, december 29, 2020 january 12, 2021. Morbidity and Mortality Weekly Report, 70:95–99, 2021.

2. Nicholas G. Davies, Sam Abbott, Rosanna C. Barnard, Christopher I. Jarvis, Adam J. Kucharski, James D. Munday, Carl A. B. Pearson, Timothy W. Russell, Damien C. Tully, Alex D. Washburne, Tom Wenseleers, Amy Gimma, William Waites, Kerry L. M. Wong, Kevin van Zandvoort, Justin D. Silverman, Karla Diaz-Ordaz, Ruth Keogh, Rosalind M. Eggo, Sebastian Funk, Mark Jit, Katherine E. Atkins, and W. John Edmunds. Estimated transmissibility and impact of sars-cov-2 lineage b.1.1.7 in england. Science, 372(6538), 2021.

3. Delphine Planas, Timothée Bruel, Ludivine Grzelak, Florence Guivel-Benhassine, Isabelle Staropoli, Françoise Porrot, Cyril Planchais, Julian Buchrieser, Maaran Michael Rajah, Elodie Bishop, Mélanie Albert, Flora Donati, Matthieu Prot, Sylvie Behillil, Vincent Enouf, Marianne Maquart, Mounira Smati-Lafarge, Emmanuelle Varon, Frédérique Schortgen, Layla Yahyaoui, Maria Gonzalez Jérôme D. Sèze, Hélène Péré, David Veyer, Aymeric Sève, Etienne Simon-Lorière, Samira Fafi-Kremer, Karl Stefic, Hugo Mouquet, Laurent Hocqueloux, Sylvie van der Werf, Thierry Prazuck, and Olivier Schwartz. Sensitivity of infectious sars-cov-2 b.1.1.7 and b.1.351 variants to neutralizing antibodies. Nature Medicine, 27(5):917–924, May 2021.

4. Alexander Muik, Ann-Kathrin Wallisch, Bianca Sänger, Kena A. Swanson, Julia Mühl, Wei Chen, Hui Cai, Daniel Maurus, Ritu Sarkar, Özlem Türeci, Philip R. Dormitzer, and Uğur Şahin. Neutralization of sars-cov-2 lineage b.1.1.7 pseudovirus by bnt162b2 vaccine–elicited human sera. Science, 371(6534):1152–1153, 2021.

5. Xiaoying Shen, Haili Tang, Charlene McDanal, Kshitij Wagh, William Fischer, James Theiler, Hyejin Yoon, Dapeng Li, Barton F. Haynes, Kevin O. Sanders, Sandrasegaram Gnanakaran, Nick Hengartner, Rolando Pajon, Gale Smith, Gregory M. Glenn, Bette Korber, and David C. Montefiori. Sars-cov-2 variant b.1.1.7 is susceptible to neutralizing antibodies elicited by ancestral spike vaccines. Cell Host & Microbe, 29(4):529–539.e3, 2021.

6. Matthew McCallum, Jessica Bassi, Anna De Marco, Alex Chen, Alexandra C. Walls, Julia Di Iulio, M. Alejandra Tortorici, Mary-Jane Navarro, Chiara Silacci-Fregni, Christian Saliba, Maria Agostini, Dora Pinto, Katja Culap, Siro Bianchi, Stefano Jaconi, Elisabetta Cameroni, John E. Bowen, Sasha W Tilles, Matteo Samuele Pizzuto, Sonja Bernasconi Guastalla, Giovanni Bona, Alessandra Franzetti Pellanda, Christian Garzoni, Wesley C. Van Voorhis, Laura E. Rosen, Gyorgy Snell, Amalio Telenti, Herbert W. Virgin, Luca Piccoli, Davide Corti, and David Veesler. Sars-cov-2 immune evasion by variant b.1.427/b.1.429. bioRxiv, 2021.

7. Abdul Aleem, Abdul Bari Akbar Samad, and Amy K. Slenker. Emerging Variants of SARS-CoV-2 And Novel Therapeutics Against Coronavirus (COVID-19). StatPearls Publishing, Treasure Island (FL), 2021.

8. Erica Lasek-Nesselquist, Pascal Lapierre, Erasmus Schneider, Kirsten St. George, and Janice Pata. The localized rise of a b.1.526 sars-cov-2 variant containing an e484k mutation in new york state. medRxiv, 2021.

9. Hao Zhou, Belinda M. Dcosta, Marie I. Samanovic, Mark J. Mulligan, Nathaniel R. Landau, and Takuya Tada. B.1.526 sars-cov-2 variants identified in new york city are neutralized by vaccine-elicited and therapeutic monoclonal antibodies. bioRxiv, 2021.

10. Manar Alkuzweny Marco Cano Emily Haag Jerry Zhou Mark Zeller Nate Matteson Kristian G. Andersen Chunlei Wu Andrew I. Su Karthik Gangavarapu Laura D. Hughes Julia L. Mullen Ginger Tsueng Alaa Abdel Latif and the Center for Viral Systems Biology. outbreak.info.

11. Stefan Elbe and Gemma Buckland-Merrett. Data, disease and diplomacy: Gisaid’s innovative contribution to global health. Global Challenges, 1(1):33–46, 2017.

12. Ensheng Dong, Hongru Du, and Lauren Gardner. An interactive web-based dashboard to track covid-19 in real time. The Lancet Infectious Diseases, 20(5):533–534, 2020.

13. U.s. census bureau. state population totals: 2010-2019.

14. Wes McKinney et al. Data structures for statistical computing in python. In Proceedings of the 9th Python in Science Conference, volume 445, pages 51–56. Austin, TX, 2010.

15. Pei Sen, Teresa K Yamana, Sasikiran Kandula, Marta Galanti, and Jeffrey Shaman. Burden and characteristics of COVID-19 in the united states during 2020. Nature, 598(7880):338–341, August 2021.

16. Ulrike Groemping. Relative importance for linear regression in r: The package relaimpo. Journal of Statistical Software, Articles, 17(1):1–27, 2006.

17. Corinne N. Thompson, Scott Hughes, Stephanie Ngai, Jennifer Baumgartner, Jade C. Wang, Emily McGibbon, Katelynn Devinney, Elizabeth Luoma, Daniel Bertolino, Christina Hwang, Kelsey Kepler, Cybill Del Castillo, Melissa Hopkins, Henry Lee, Andrea K. DeVito, Jennifer L. Rakeman, PhD1, and Anne D. Fine. Rapid emergence and epidemiologic characteristics of the sars-cov-2 b.1.526 variant - new york city, new york, january 1-april 5, 2021. MMWR. Morbidity and mortality weekly report, 70(19):712–716, May 2021. 33983915[pmid].

